# Investigation of a SARS-CoV-2 outbreak in a Texas summer camp resulting from a single introduction

**DOI:** 10.1101/2022.05.29.22275277

**Authors:** Daniele M. Swetnam, R. Elias. Alvarado, Stephanea Sotcheff, Brooke M. Mitchell, Allan McConnell, Rafael R.G. Machado, Nehad Saada, Florence P. Haseltine, Sara Maknojia, Anajane Smith, Ping Ren, Philip Keiser, Scott C. Weaver, Andrew Routh

**Author notes:** **Address for Correspondence** Daniele Swetnam, Andrew Routh.

## Abstract

SARS-CoV-2 is the etiological agent responsible for the COVID-19 pandemic. It is estimated that only 10 aerosol-borne virus particles are sufficient to establish a secondary infection with SARS-CoV-2. However, the dispersal pattern of SARS-CoV-2 is highly variable and only 10– 20% of cases are responsible for up 80% of secondary infections. The heterogeneous nature of SARS-CoV-2 transmission suggests that super-spreader events play an important role in viral transmission. Super-spreader events occur when a single person is responsible for an unusually high number of secondary infections due to a combination of biological, environmental, and/or behavioral factors. While super-spreader events have been identified as a significant factor driving SARS-CoV-2 transmission, epidemiologic studies have consistently shown that education settings do not play a major role in community transmission. However, an outbreak of SARS-CoV-2 was recently reported among 186 children (aged 10-17) and adults (aged 18 +) after attending an overnight summer camp in Texas in June 2021. To understand the transmission dynamics of the outbreak, RNA was isolated from 36 nasopharyngeal swabs collected from patients that attended the camp and 19 control patients with no known connection to the outbreak. Genome sequencing on the Oxford Nanopore platform was performed using the ARTIC approaches for library preparation and bioinformatic analysis. SARS-CoV-2 amplicons were produced from all RNA samples and >70% of the viral genome was successfully reconstructed with >10X coverage for 46 samples. Phylogenetic methods were used to estimate the transmission history and suggested that the outbreak was the result of a single introduction. We also found evidence for secondary transmission from campers to the community. Together, these findings demonstrate that super-spreader events may occur during large gatherings of children.

## Main Text

The rapid spread of Severe Acute Respiratory Syndrome Coronavirus 2 (SARS-CoV-2), the virus driving the Coronavirus disease 2019 (COVID-19) pandemic, is partly due to the highly infectious nature of the virus. Approximately 10 aerosol-borne virus particles are sufficient to establish a novel infection (1). However, the dispersal pattern (K) of SARS-CoV-2 is highly variable. Approximately 60–75% of infected patients do not transmit SARS-CoV-2 infection, and only 15% of cases are responsible for up to 80% of secondary infections (2). The heterogenoeus dispersal dynamics of SARS-CoV-2 suggest that super-spreader events play a major role in transmission dynamics.

Super-spreader events result in the transmission of SARS-CoV-2 from an infected individual to an unusually high number of persons due to some combination of environmental, biological and/or behavioral factors. Such events have been reported frequently, especially involving mass gatherings (reviewed (3)). Given the well-documented role of super-spreader events in the transmission of SARS-CoV-2, there is considerable concern over the risk of super-spreader events occurring in educational settings among children, their caregivers, and staff. Studies in Europe (4–11), North America (12–15), and Oman (16) have repeatedly shown that educational settings are not major drivers of SARS-CoV-2 transmission in the community, despite the dense populations and prolonged exposure of children and adolescents in classrooms. Even when infected, children usually develop milder disease (17) and emit fewer virus particles than adults (18). Outbreaks among schools report low secondary attack rates (4,19) with child-to-child transmission, which is significantly lower than adult-to-adult and adult-to-child transmission (19–22). However, several studies have noted an important distinction between transmission among young children (age 6-12) and adolescents (23,24) with reports of larger and more frequent outbreaks among secondary schools (25). Rather, household transmission appears to be a more important driver of SARS-CoV-2 spread (14,22,26).

While educational settings have not been a major source of SARS-CoV-2 transmission, large outbreaks among children and adolescents have been reported in several summer camps throughout the United States of America (27–29), with primary attack rates as high as 46% (28) in Georgia and even 76% in Wisconsin (29). However, some reports have demonstrated success preventing and event halting the transmission of SARS-CoV-2 with multi-layered mitigation approaches, including but not limited to vaccination, pre-arrival quarantine, pre-post arrival testing, masking, and social distancing (30–32). However, studies investigating SARS-CoV-2 outbreaks in summer camps have relied on diagnostic and epidemiological methods alone and have lacked phylogenetic approaches that could provide insights into the patterns of viral transmission.

Such an outbreak occurred at an overnight summer camp in Texas during June 2021. Four hundred and fifty-one individuals attended including 364 youths (ages 10-17) and 87 adults (age 18 or older). At the conclusion of the camp, attendees began to show symptoms and tested positive for SARS-CoV-2 by RT-PCR. An investigation of the outbreak led to the identification of 186 SARS-CoV-2 positive cases (33). The vaccination rate among attendees was low (19% fully vaccinated and 6% partially vaccinated), pre-arrival testing was not required for camp attendance, and no post-arrival testing was conducted. The primary attack rate within the camp was 41% (48% among unvaccinated attendees and 20% among vaccinated), which is consistent with outbreaks reported in overnight summer camps in Georgia (28).

Notably, the Texas outbreak occurred at the beginning of the Delta variant wave in the US, while cases were still low in the community but rapidly increasing. To better understand the pattern of viral transmission, we obtained 55 SARS-CoV-2 positive nasopharyngeal swabs collected by the University of Texas Medical Branch, Galveston, TX from patients who attended the summer camp (36) as well as from unrelated community members who tested positive during the same time period (17). RNA was extracted from the swabs as previously described (34) and used for deep sequencing with the well-defined ARTIC nanopore approach and associated bioinformatic pipeline (34). SARS-CoV-2 reads were detected in all samples, and 70% genome coverage greater than 10X was achieved for 43 samples (31 camp attendees and 13 Galveston County residents). The results were aligned with 3 additional genomes that were isolated from an Arlington, TX family that tested positive following contact with asymptomatic attendees after the camp concluded. The Arlington genomes were sequenced using similar methods (35). All genomes are available on GSAID EPI_ISL_12486186-213 (Table 1).

**Table 1.** Sequence Metadata. All relevant metadata are provided including sample names, dates the samples were collected, type of patients (camper or community member) and the GSAID accession number.

| Sample Name | Collection Date | Patient Type | Accession Number |
| --- | --- | --- | --- |
| MicroGNL-1131 | 6/28/21 | Camper | EPI_ISL_12486192 |
| MicroGNL-1134 | 6/28/21 | Community Member | EPI_ISL_12486173 |
| MicroGNL-1136 | 6/28/21 | Camper | EPI_ISL_12486193 |
| MicroGNL-1138 | 6/28/21 | Camper | EPI_ISL_12486179 |
| MicroGNL-1137 | 6/29/21 | Community Member | EPI_ISL_12740992 |
| MicroGNL-1139 | 6/28/21 | Camper | EPI_ISL_12486194 |
| MicroGNL-1140 | 6/28/21 | Community Member | EPI_ISL_12486174 |
| MicroGNL-1149 | 6/29/21 | Camper | EPI_ISL_12486180 |
| MicroGNL-1150 | 6/29/21 | Camper | EPI_ISL_12486181 |
| MicroGNL-1151 | 6/29/21 | Camper | EPI_ISL_12486182 |
| MicroGNL-1152 | 6/29/21 | Camper | EPI_ISL_12486183 |
| MicroGNL-1153 | 6/28/21 | Community Member | EPI_ISL_12486168 |
| MicroGNL-1154 | 6/29/21 | Community Member | EPI_ISL_12486184 |
| MicroGNL-1155 | 6/28/21 | Camper | EPI_ISL_12486195 |
| MicroGNL-1156 | 6/29/21 | Camper | EPI_ISL_12486185 |
| MicroGNL-1157 | 6/29/21 | Camper | EPI_ISL_12486186 |
| MicroGNL-1160 | 6/29/21 | Camper | EPI_ISL_12486187 |
| MicroGNL-1161 | 6/29/21 | Community Member | EPI_ISL_12486172 |
| MicroGNL-1163 | 6/29/21 | Community Member | EPI_ISL_12486175 |
| MicroGNL-1164 | 6/30/21 | Camper | EPI_ISL_12486196 |
| MicroGNL-1166 | 6/30/21 | Community Member | EPI_ISL_12486176 |
| MicroGNL-1167 | 6/29/21 | Camper | EPI_ISL_12486197 |
| MicroGNL-1168 | 6/29/21 | Camper | EPI_ISL_12486198 |
| MicroGNL-1170 | 6/30/21 | Camper | EPI_ISL_12486199 |
| MicroGNL-1171 | 6/29/21 | Camper | EPI_ISL_12486188 |
| MicroGNL-1172 | 6/30/21 | Camper | EPI_ISL_12486200 |
| MicroGNL-1173 | 6/30/21 | Community Member | EPI_ISL_12486177 |
| MicroGNL-1174 | 6/29/21 | Camper | EPI_ISL_12486189 |
| MicroGNL-1175 | 6/29/21 | Camper | EPI_ISL_12486201 |
| MicroGNL-1180 | 6/30/21 | Camper | EPI_ISL_12486202 |
| MicroGNL-1182 | 6/30/21 | Camper | EPI_ISL_12486203 |
| MicroGNL-1185 | 6/30/21 | Camper | EPI_ISL_12486204 |
| MicroGNL-1188 | 7/3/21 | Camper | EPI_ISL_12486190 |
| MicroGNL-1189 | 7/4/21 | Community Member | EPI_ISL_12486178 |
| MicroGNL-1192 | 7/2/21 | Camper | EPI_ISL_12486205 |
| MicroGNL-1199 | 7/4/21 | Camper | EPI_ISL_12486206 |
| MicroGNL-1201 | 7/4/21 | Camper | EPI_ISL_12486207 |
| MicroGNL-1204 | 7/6/21 | Community Member | EPI_ISL_12486169 |
| MicroGNL-1205 | 7/6/21 | Camper | EPI_ISL_12486191 |
| MicroGNL-1211 | 7/6/21 | Community Member | EPI_ISL_12486170 |
| MicroGNL-1213 | 7/5/21 | Camper | EPI_ISL_12486208 |
| MicroGNL-1217 | 7/7/21 | Community Member | EPI_ISL_12486171 |
| MicroGNL-1219 | 7/7/21 | Camper | EPI_ISL_12486209 |
| MicroGNL-1223 | 7/7/21 | Camper | EPI_ISL_12486210 |
| NTD_14603 | 7/7/21 | Community Member | EPI_ISL_12486211 |
| NTW_14608 | 7/7/21 | Community Member | EPI_ISL_12486212 |
| NTF_14607 | 7/7/21 | Community Member | EPI_ISL_12486213 |
| NTS_14602 | 7/7/21 | Community Member | EPI_ISL_12740993 |

The evolutionary history of the genomes was inferred using a maximum likelihood approach implemented with IQ-Tree (36) (Figure 1A). The phylogenetic tree demonstrated that all the SARS-CoV-2 genomes collected from camp attendees shared a single common ancestor, including the genomes collected from the family from Arlington, TX that became sick after contact with camp attendees. This suggests that the outbreak was initiated from a single infected individual. Genomes from campers that shared suspected risk factors, including bus or cabin assignment, did not cluster together. Similarly, the genomes collected from siblings did not appear to cluster together, suggesting that transmission occurred while the youths were at the camp as opposed to when they returned home. However, these negative results should be interpreted with caution because there was insufficient diversity to resolve all branches within the phylogeny. The lack of diversity also prevented generation of a transmission tree, which may have provided additional epidemiologically relevant insights. This was unsurprising, given the short duration of the outbreak (5 days) relative to the generation time (delta variant mean=4.7 days, 95% CI=4.1-5.6 days)(37). Interestingly, several genomes from patients that did not attend the camp clustered among the campers, suggesting that transmission occurred from campers to the community.

**Figure 1.**
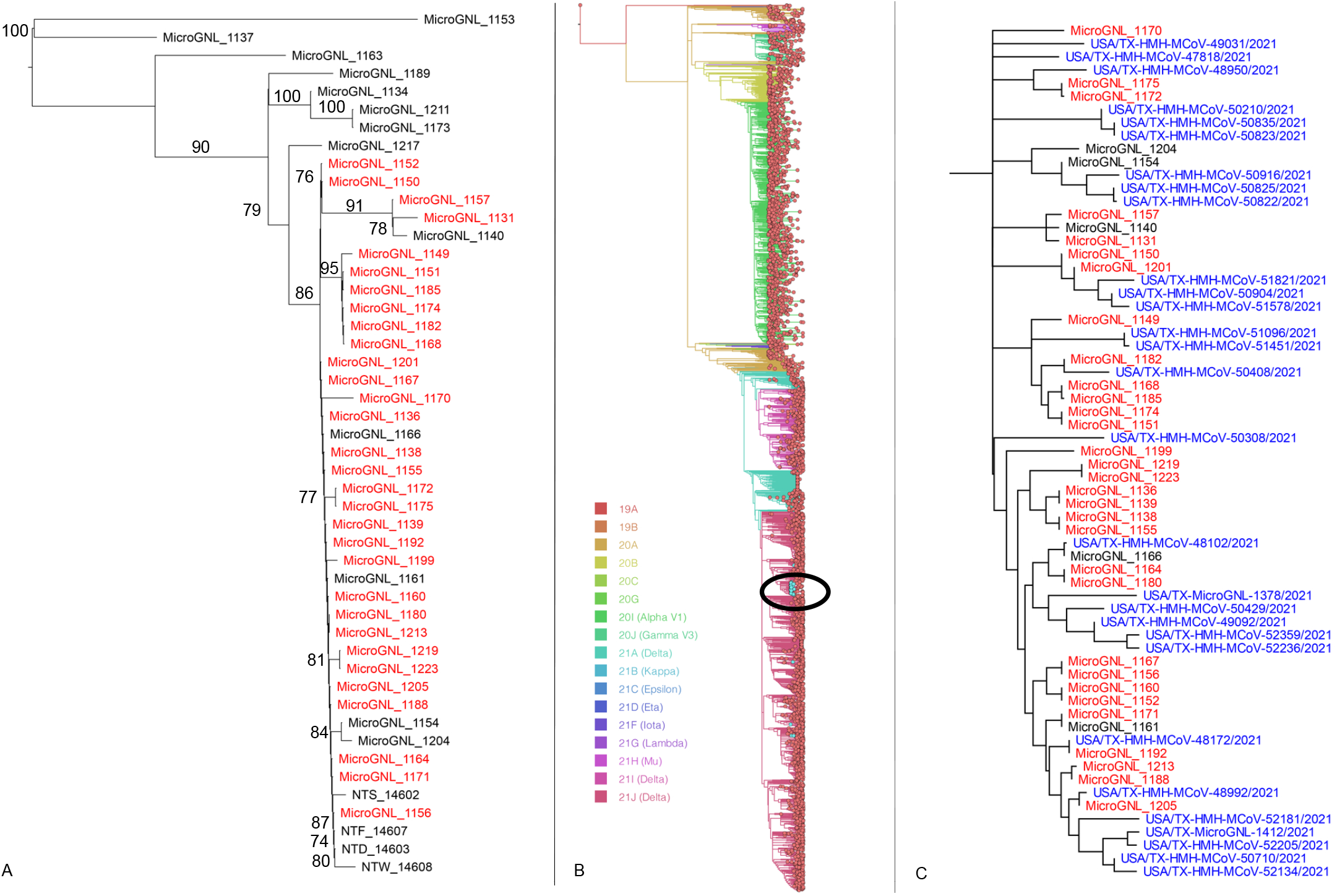
Evolutionary history of genomes collected from camp attendees. The genomes collected from camp attendees with at least 70% genomic coverage were aligned. (A) The evolutionary history was inferred with IQ-Tree using the Maximum likelihood method based on the general time reversible model. A discrete gamma distribution was used to model variation among sites and allow for invariable sites. The consensus tree following 1000 UF bootstraps is shown. Genomes collected from camp attendees are shown in red and genomes collected from community members are shown in black. Bootstrap values greater than 70 are provided. (B) All complete genomes (n=4085) available on GSAID collected in Harris and Galveston Counties between May and July were combined with the genomes sequenced in this study. The evolutionary history was inferred using the Nextstrain platform, which using Augar to preform bioinformatic analysis including aligning and filtering the genomes, generating a phylogeny with IQ-Tree, and removing polytomies, inferring node dates, and pruning branches with TimeTree. The phylogeny was visualized with Auspice and Figtree. The branches of the phylogeny were colored to indicate clade membership. The cluster containing genomes collected from campers is indicated by the black circle and expanded (C). Genomes isolated from campers and community members are depicted in red and black, respectively. Genomes obtained from GSAID are depicted in blue.

To further investigate the role of the camp outbreak in community transmission, all complete genomes published on GISAID collected in Galveston County and Harris County, Texas (4085 genomes in total) were compared to the genomes collected from the campers. Phylogenetic analysis using the Nextstrain platform (38) identified 29 genomes that clustered among the camper’s genomes that were collected after the camp concluded between June 28, 2021 and July 30, 2021, suggesting that community transmission originating from the camp outbreak continued at least until the end of July (Figure 1B-C).

It is essential to understand the mechanisms that support super-spreader events, such as the Texas summer camp outbreak of 2021, and the risk factors associated with them. Taken together, our study demonstrates that the Texas camp outbreak was likely the result of a single introduction that spread in the camp environment and eventually into the community, creating a chain of transmission that persisted until at least July 30, 2021. This study also highlights the risks associated with overnight summer camps that do not employ adequate prevention strategies, such vaccination, pre- and post-arrival testing, etc. Furthermore, this study is the first of our knowledge to combine epidemiological, genomic, and phylogenetic approaches to investigate an outbreak at an overnight camp. It illustrates the importance of multidisciplinary collaborations between public health specialists and evolutionary virologist in responding to this and future pandemics.

## Data Availability

All data produced in the present study are available upon reasonable request to the authors.

## Acknowledgments

The authors of this manuscript gratefully acknowledge the following funding sources: ‘Data collection grant’ and ‘COVID-19 funding’ from IHII to ALR; NIH grant R21AI151725 from to ALR; NIH grant R24 AI120942 to SW; The Sealy and Smith Foundation to SW and UTMB; CDC Contract 200-2021-11195 to ALR and Kempner Fellowship to DMS; FAPESP Fellowship 2019/27803 to RRGM, the West African Center for Emerging Infectious Diseases to SW, BM, AM, RM, NS.

## References

1. Bazant MZ, Bush JWM. A guideline to limit indoor airborne transmission of COVID-19. [cited 2022 Mar 25]; Available from: https://doi.org/10.1073/pnas.2018995118

2. Sun K, Wang W, Gao L, Wang Y, Luo K, Ren L, et al. Transmission heterogeneities, kinetics, and controllability of SARS-CoV-2. Science (80-) [Internet]. 2021 Jan 15 [cited 2022 Mar 25];371(6526). Available from: https://www.science.org/doi/full/10.1126/science.abe2424

3. Majra D, Benson J, Pitts J, Stebbing J. SARS-CoV-2 (COVID-19) superspreader events. 2021 Jan 1 [cited 2022 Mar 25];82(1). Available from: https://doi.org/10.1016/j.jinf.2020.11.021

4. Powell AA, Ireland G, Baawuah F, Beckmann J, Okike IO, Ahmad S, et al. Secondary attack rates in primary and secondary school bubbles following a confirmed case: Active, prospective national surveillance, November to December 2020, England. PLoS One [Internet]. 2022 Feb 1 [cited 2022 Mar 4];17(2). Available from: https://pubmed.ncbi.nlm.nih.gov/35171942/

5. Young BC, Eyre DW, Kendrick S, White C, Smith S, Beveridge G, et al. Daily testing for contacts of individuals with SARS-CoV-2 infection and attendance and SARS-CoV-2 transmission in English secondary schools and colleges: an open-label, cluster-randomised trial. Lancet (London, England) [Internet]. 2021 Oct 2 [cited 2022 Mar 9];398(10307):1217–29. Available from: https://pubmed.ncbi.nlm.nih.gov/34534517/

6. Mensah AA, Sinnathamby M, Zaidi A, Coughlan L, Simmons R, Ismail SA, et al. SARS-CoV-2 infections in children following the full re-opening of schools and the impact of national lockdown: Prospective, national observational cohort surveillance, July-December 2020, England. J Infect [Internet]. 2021 Apr 1 [cited 2022 Mar 4];82(4):67–74. Available from: https://pubmed.ncbi.nlm.nih.gov/33639175/

7. White P, O’Sullivan MB, Murphy N, Stapleton J, Dillon A, Brennan A, et al. An investigation into the rates of transmission of SARS-CoV-2 during the first 6 weeks of the 2020-2021 academic year in primary and post-primary schools in Cork and Kerry, Ireland. Ir J Med Sci [Internet]. 2021 [cited 2022 Mar 4]; Available from: https://pubmed.ncbi.nlm.nih.gov/33792855/

8. Gandini S, Rainisio M, Iannuzzo ML, Bellerba F, Cecconi F, Scorrano L. A cross-sectional and prospective cohort study of the role of schools in the SARS-CoV-2 second wave in Italy. Lancet Reg Heal -Eur [Internet]. 2021 Jun 1 [cited 2022 Mar 10];5. Available from: http://www.thelancet.com/article/S2666776221000697/fulltext

9. Ulyte A, Radtke T, Abela IA, Haile SR, Berger C, Huber M, et al. Clustering and longitudinal change in SARS-CoV-2 seroprevalence in school children in the canton of Zurich, Switzerland: prospective cohort study of 55 schools. BMJ [Internet]. 2021 Mar 17 [cited 2022 Mar 4];372. Available from: https://pubmed.ncbi.nlm.nih.gov/33731327/

10. Macartney K, Quinn HE, Pillsbury AJ, Koirala A, Deng L, Winkler N, et al. Transmission of SARS-CoV-2 in Australian educational settings: a prospective cohort study. Lancet Child Adolesc Heal [Internet]. 2020 Nov 1 [cited 2022 Mar 9];4(11):807–16. Available from: https://pubmed.ncbi.nlm.nih.gov/32758454/

11. Brandal LT, Ofitserova TS, Meijerink H, Rykkvin R, Lund HM, Hungnes O, et al. Minimal transmission of SARS-CoV-2 from paediatric COVID-19 cases in primary schools, Norway, August to November 2020. Euro Surveill [Internet]. 2021 Dec 1 [cited 2022 Mar 9];26(1). Available from: https://pubmed.ncbi.nlm.nih.gov/33413743/

12. Doyle T, Kendrick K, Troelstrup T, Gumke M, Edwards J, Chapman S, et al. COVID-19 in Primary and Secondary School Settings During the First Semester of School Reopening -Florida, August-December 2020. MMWR Morb Mortal Wkly Rep [Internet]. 2021 [cited 2022 Mar 4];70(12):437–41. Available from: https://pubmed.ncbi.nlm.nih.gov/33764962/

13. Hershow RB, Wu K, Lewis NM, Milne AT, Currie D, Smith AR, et al. Low SARS-CoV-2 Transmission in Elementary Schools - Salt Lake County, Utah, December 3, 2020-January 31, 2021. MMWR Morb Mortal Wkly Rep [Internet]. 2021 [cited 2022 Mar 4];70(12):442– 8. Available from: https://pubmed.ncbi.nlm.nih.gov/33764967/

14. Peetluk LS, Rebeiro PF, Edwards KM, Banerjee R, Mallal SA, Aronoff DM, et al. Minimal in-school SARS-CoV-2 transmission with strict mitigation protocols at two independent schools in Nashville, TN. medRxiv Prepr Serv Heal Sci [Internet]. 2021 Nov 10 [cited 2022 Mar 9]; Available from: https://pubmed.ncbi.nlm.nih.gov/34790987/

15. Bark D, Dhillon N, St-Jean M, Kinniburgh B, McKee G, Choi A. SARS-CoV-2 transmission in kindergarten to grade 12 schools in the Vancouver Coastal Health region: a descriptive epidemiologic study. C open [Internet]. 2021 Jul 1 [cited 2022 Mar 9];9(3):E810–7. Available from: https://pubmed.ncbi.nlm.nih.gov/34429325/

16. Alqayoudhi A, Al Manji A, Al khalili S, Al Maani A, Alkindi H, Alyaquobi F, et al. The role of children and adolescents in the transmission of SARS-CoV-2 virus within family clusters: A large population study from Oman. J Infect Public Health [Internet]. 2021 Nov 1 [cited 2022 Mar 9];14(11):1590–4. Available from: https://pubmed.ncbi.nlm.nih.gov/34627056/

17. Ludvigsson JF. Systematic review of COVID-19 in children shows milder cases and a better prognosis than adults. Acta Paediatr [Internet]. 2020 Jun 1 [cited 2022 Mar 10];109(6):1088–95. Available from: https://pubmed.ncbi.nlm.nih.gov/32202343/

18. Fleischer M, Schumann L, Hartmann A, Walker RS, Ifrim L, Zadow D von, et al. Pre-adolescent children exhibit lower aerosol particle volume emissions than adults for breathing, speaking, singing and shouting. J R Soc Interface [Internet]. 2022 Feb 23 [cited 2022 Mar 4];19(187). Available from: /pmc/articles/PMC8864358/

19. Aizawa Y, Shobugawa Y, Tomiyama N, Nakayama H, Takahashi M, Yanagiya J, et al. Coronavirus Disease 2019 Cluster Originating in a Primary School Teachers’ Room in Japan. Pediatr Infect Dis J [Internet]. 2021 [cited 2022 Mar 9];40(11):E418–23. Available from: https://pubmed.ncbi.nlm.nih.gov/34561385/

20. Gold JAW, Gettings JR, Kimball A, Franklin R, Rivera G, Morris E, et al. Clusters of SARS-CoV-2 Infection Among Elementary School Educators and Students in One School District -Georgia, December 2020-January 2021. MMWR Morb Mortal Wkly Rep [Internet]. 2021 [cited 2022 Mar 4];70(8):289–92. Available from: https://pubmed.ncbi.nlm.nih.gov/33630823/

21. Jang J, Hwang MJ, Kim YY, Park SY, Yoo M, Kim SS, et al. Epidemiological Characteristics and Transmission Patterns of COVID-19 Cases Among Children and Adolescents Aged 0-18 Years in South Korea. Risk Manag Healthc Policy [Internet]. 2022 [cited 2022 Mar 9];15:219–27. Available from: https://pubmed.ncbi.nlm.nih.gov/35173498/

22. Stich M, Elling R, Renk H, Janda A, Garbade SF, Müller B, et al. Transmission of Severe Acute Respiratory Syndrome Coronavirus 2 in Households with Children, Southwest Germany, May-August 2020. Emerg Infect Dis [Internet]. 2021 Dec 1 [cited 2022 Mar 9];27(12):3009–19. Available from: https://pubmed.ncbi.nlm.nih.gov/34695369/

23. Boey L, Roelants M, Merckx J, Hens N, Desombere I, Duysburgh E, et al. Age-dependent seroprevalence of SARS-CoV-2 antibodies in school-aged children from areas with low and high community transmission. Eur J Pediatr [Internet]. 2022 Feb 1 [cited 2022 Mar 10];181(2):571–8. Available from: https://pubmed.ncbi.nlm.nih.gov/34455523/

24. Irfan O, Li J, Tang K, Wang Z, Bhutta ZA. Risk of infection and transmission of SARS-CoV-2 among children and adolescents in households, communities and educational settings: A systematic review and meta-analysis. J Glob Health [Internet]. 2021 [cited 2022 Mar 9];11:1–15. Available from: https://pubmed.ncbi.nlm.nih.gov/34326997/

25. Aiano F, Mensah AA, McOwat K, Obi C, Vusirikala A, Powell AA, et al. COVID-19 outbreaks following full reopening of primary and secondary schools in England: Cross-sectional national surveillance, November 2020. Lancet Reg Heal Eur [Internet]. 2021 Jul 1 [cited 2022 Mar 9];6. Available from: https://pubmed.ncbi.nlm.nih.gov/34278370/

26. Viner R, Waddington C, Mytton O, Booy R, Cruz J, Ward J, et al. Transmission of SARS-CoV-2 by children and young people in households and schools: A meta-analysis of population-based and contact-tracing studies. J Infect [Internet]. 2022 Mar 1 [cited 2022 Mar 10];84(3):361. Available from: /pmc/articles/PMC8694793/

27. Tonzel JL, Sokol T. COVID-19 Outbreaks at Youth Summer Camps — Louisiana, June–July 2021. Morb Mortal Wkly Rep [Internet]. 2021 [cited 2022 Mar 10];70(40):1425. Available from: /pmc/articles/PMC8519274/

28. Szablewski CM, Chang KT, Brown MM, Chu VT, Yousaf AR, Anyalechi N, et al. SARS-CoV-2 Transmission and Infection Among Attendees of an Overnight Camp — Georgia, June 2020. Morb Mortal Wkly Rep [Internet]. 2020 Aug 7 [cited 2022 Mar 10];69(31):1023. Available from: /pmc/articles/PMC7454898/

29. Pray IW, Gibbons-Burgener SN, Rosenberg AZ, Cole D, Borenstein S, Bateman A, et al. COVID-19 Outbreak at an Overnight Summer School Retreat - Wisconsin, July-August 2020. MMWR Morb Mortal Wkly Rep [Internet]. 2020 Oct 30 [cited 2022 Mar 10];69(43):1600–4. Available from: https://pubmed.ncbi.nlm.nih.gov/33119558/

30. Kowalsky RH, Fine S, Eisenberg M. Exclusion of SARS-COV-2 From Two Maine Overnight Camps July-August 2020. Disaster Med Public Health Prep [Internet]. 2021 [cited 2022 Mar 10];1–10. Available from: https://pubmed.ncbi.nlm.nih.gov/33762061/

31. Blaisdell LL, Cohn W, Pavell JR, Rubin DS, Vergales JE. Preventing and Mitigating SARS-CoV-2 Transmission — Four Overnight Camps, Maine, June–August 2020. MMWR Morb Mortal Wkly Rep. 2020 Sep 4;69(35):1216–20.

32. Braun KVN, Drexler M, Rozenfeld RA, Deener-Agus E, Greenstein R, Agus M, et al. Multicomponent Strategies to Prevent SARS-CoV-2 Transmission -Nine Overnight Youth Summer Camps, United States, June-August 2021. MMWR Morb Mortal Wkly Rep [Internet]. 2021 [cited 2022 Mar 10];70(40):1420–4. Available from: https://pubmed.ncbi.nlm.nih.gov/34618796/

33. Baker JM, Shah MM;, O’Hegarty M, Pomeroy M, Keiser P, Ren P, et al. Primary and Secondary Attack Rates by Vaccination Status after a SARS-CoV-2 B.1.617.2 (Delta) Variant Outbreak at a Youth Summer Camp — Texas, June 2021. submitted.

34. Jaworski E, Langsjoen RM, Mitchell B, Judy B, Newman P, Plante JA, et al. Tiled-clickseq for targeted sequencing of complete coronavirus genomes with simultaneous capture of rna recombination and minority variants. Elife. 2021 Sep 1;10.

35. Zhao LP, Roychoudhury P, Gilbert P, Schiffer J, Lybrand TP, Payne TH, et al. Mutations in viral nucleocapsid protein and endoRNase are discovered to associate with COVID19 hospitalization risk. Sci Reports 2022 121 [Internet]. 2022 Jan 24 [cited 2022 Apr 18];12(1):1–11. Available from: https://www.nature.com/articles/s41598-021-04376-4

36. Nguyen LT, Schmidt HA, Von Haeseler A, Minh BQ. IQ-TREE: A Fast and Effective Stochastic Algorithm for Estimating Maximum-Likelihood Phylogenies. Mol Biol Evol [Internet]. 2015 Jan 1 [cited 2022 Apr 14];32(1):268. Available from: /pmc/articles/PMC4271533/

37. Hart WS, Miller E, Andrews NJ, Waight P, Maini PK, Funk S, et al. Generation time of the alpha and delta SARS-CoV-2 variants: an epidemiological analysis. Lancet Infect Dis [Internet]. 2022 Feb [cited 2022 Mar 25];0(0). Available from: http://www.thelancet.com/article/S1473309922000019/fulltext

38. Hadfield J, Megill C, Bell SM, Huddleston J, Potter B, Callender C, et al. Nextstrain: real-time tracking of pathogen evolution. Kelso J, editor. Bioinformatics [Internet]. 2018 Dec 1 [cited 2019 Feb 15];34(23):4121–3. Available from: https://academic.oup.com/bioinformatics/article/34/23/4121/5001388

